# Inflight Transmission of COVID-19 Based on Aerosol Dispersion Data

**DOI:** 10.1101/2021.01.08.21249439

**Authors:** Zhaozhi Wang, Edwin R. Galea, Angus Grandison, John Ewer, Fuchen Jia

**Affiliations:** Fire Safety Engineering Group, Faculty of Liberal Arts and Sciences, University of Greenwich, London UK

**Author notes:** To whom correspondence should be addressed: E.R.Galea, Fire Safety Engineering Group, University of Greenwich, 30 Park Row, Greenwich, London SE10 9LS, UK, EMAIL.

**Keywords:** SARS-CoV-2, Wells-Riley model, Inflight transmission, Infection probability, Quanta, COVID-19

## Abstract

**Background:** An issue of concern to the travelling public is the possibility of in-flight transmission of COVID-19 during long- and short-haul flights. The aviation industry maintain the probability of contracting the illness is small based on reported cases, modelling and data from aerosol dispersion experiments conducted on-board aircraft.

**Methods:** Using experimentally derived aerosol dispersion data for a B777-200 aircraft and a modified version of the Wells-Riley equation we estimate inflight infection probability for a range of scenarios involving quanta generation rate and face mask efficiency. Quanta generation rates were selected based on COVID-19 events reported in the literature while mask efficiency was determined from the aerosol dispersion experiments.

**Results:** The MID-AFT cabin exhibits the highest infection probability. The calculated maximum individual infection probability (without masks) for a 2-hour flight in this section varies from 4.5% for the “Mild Scenario” to 60.2% for the “Severe Scenario” although the corresponding average infection probability varies from 0.1% to 2.5%. For a 12-hour flight, the corresponding maximum individual infection probability varies from 24.1% to 99.6% and the average infection probability varies from 0.8% to 10.8%. If all passengers wear face masks throughout the 12-hour flight, the average infection probability can be reduced by approximately 73%/32% for high/low efficiency masks. If face masks are worn by all passengers except during a one-hour meal service, the average infection probability is increased by 59%/8% compared to the situation where the mask is not removed.

**Conclusions:** This analysis has demonstrated that while there is a significant reduction in aerosol concentration due to the nature of the cabin ventilation and filtration system, this does not necessarily mean that there is a low probability or risk of in-flight infection. However, mask wearing, particularly high-efficiency ones, significantly reduces this risk.

## Introduction

It is generally accepted that aviation has played a key role in spreading the coronavirus disease 2019 (COVID-19) around the world resulting in the 2020 pandemic [1-4]. In the very early days of the pandemic, prior to the shutting of international boarders, the introduction of flight restrictions [5, 6] and health checks at airports, passengers infected with COVID-19 that were asymptomatic, pre-symptomatic or suffering from mild symptoms are likely to have carried, albeit unknowingly, the severe acute respiratory syndrome coronavirus 2 (SARS-CoV-2) to every corner of the world.

As our awareness of the COVID-19 disease has grown, and with the introduction of measures such as quarantining, rapid contact tracking, enhanced hygiene precautions, rapid swab testing, masking and passenger temperature checks, there is a growing desire, from both the aviation industry and the public for the normalisation of national and international business/leisure flights. At the time of writing (late November 2020) as much of the world enters the festive season marked by Thanksgiving and Christmas, more and more people are considering taking flights. Given our current knowledge of SARS-CoV-2 and the measures now in place, it is less likely that flights will be a major mechanism for the continued uncontrolled spread of COVID-19 around the world. However, another issue of concern is the possibility of contracting the disease while confined to the small space of the passenger cabin, in close proximity to other passengers, especially for extended periods of time ranging from around 2-hours for short-haul flights to 12-hours or more for long-haul flights.

SARS-CoV-2 has both significant transmission from pre-symptomatic and asymptomatic persons and the secondary cases may remain asymptomatic even with a 14-day follow-up period [7]. Therefore, a COVID-19 case confirmed days after taking a flight could have been infected before departure; at the departure/arrival airport; on the journey to/from the airport; or on the flight. As timing is so critical, confirmation of inflight SARS-CoV-2 transmission cases with a high degree of certainty are limited. Seven inflight SARS-CoV-2 transmission events from 24 January 2020 to 21 September 2020 from peer-reviewed or public health publications have been comprehensively reviewed in [7]. In one of these events, it is reported that one symptomatic passenger likely infected 15 secondary cases during a 10-hour flight [8]. A recent study suggests that, despite passengers having negative SARS-CoV-2 test results pre-flight, it is likely that inflight transmission occurred [4]. Nevertheless, the aviation industry claim that even with an infectious passenger on board, the inflight infection risk is low and cite the use on-board of high ventilation rates (up to 30 air changes per hour (ACH)) and HEPA filters as effective mitigations to aerosolised dispersion of SARS-Cov-2 [9-12].

Given the difficulties with exploring anecdotal accounts of potential inflight infection events, computational modelling [9-11, 13] and experimental techniques [14, 15] have been deployed to investigate potential inflight transmission mechanisms. One of the largest experimental studies involved more than 300 experiments using United Airlines Boeing 767-300 and 777-200 twin aisle aircraft, conducted by the United States Transportation Command (US Transcom) to simulate the transportation of aerosol particles (1 to 3 µm) from a source representing a passenger infected with SARS-CoV-2 [14, 15]. These experiments showed that the aerosol was rapidly diluted by the high cabin ACH, resulting in a minimum reduction of 99.7% and an average of 99.99% for the 40+ passenger breathing zone penetration (BZP) monitoring points measured in each section of the aircraft. A main conclusion of this study was that the aerosol exposure risk is minimal even during long-haul flights.

A fundamental limitation of the analysis presented in [15] is that simply presenting the reduction in aerosol concentration, even though it is significant, does not quantify the impact this may have on the probability of infection. The Wells-Riley model [16] provides a way to estimate infection probability in a confined ventilated space using quanta generation and ventilation rates. Marcus et al. [13] used the Wells-Riley model together with a Multi-zone Markov Model to estimate the required flight-time to cause an infection. The work concluded that the time to infection is very large provided masks are worn, and so the likelihood of inflight transmission of SARS-CoV-2 is minimal, supporting the claim made in [14, 15].

However, the use of time to infection, as an assessment criterion, is overly simplistic and liable to be misunderstood. The probability determined by the Wells-Riley model represents the proportion of the population or the probability that an individual is likely to be infected by the given inhaled quanta. Quanta, a term defined by Wells [17], suggests that if a person inhales one quanta of virus, the probability that they will be infected is 63%. Therefore, rather than simply identifying the time to infection, the probability of infection associated with an exposure time should be identified when assessing the potential for inflight transmission of SARS-CoV-2.

## Method

Using experimental data for the B777-200 aircraft [14, 15] and a modified Wells-Riley model, we estimate the inflight infection probability (assuming aerosol transmission) within the MID-AFT (economy class) and FWD (business class) sections of the aircraft. These cabin sections represent the areas with the highest and lowest experimentally derived values of maximum aerosol BZP respectively.

The Wells-Riley equation [16] is given by:

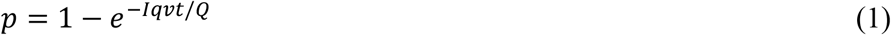

where *p* is probability of infection; *I* is number of index patients; *v* is pulmonary ventilation rate of each susceptible (m^3^/h); *Q* is space ventilation rate (m^3^/h); *q* is quanta generation rate produced by one index patient (quanta/h); and *t* is exposure time (h). The term *Iqvt*/*Q* represents the number of quanta inhaled (dose received) during the time period *t* and *Iqt* is the number of total released quanta. The experimental data [14, 15] records the percentage of released aerosols received by a susceptible, denoted as, *r*, involving a single index patient (*I* =1), which is equivalent to the ratio of *Iqvt*/*Q* to *Iqt*. Therefore, the infection probability based on the data in [14, 15] can be expressed as:

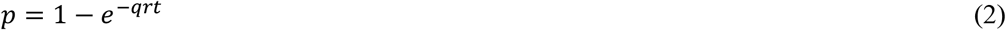

If the index patient wears a face mask with efficiency *a* in preventing aerosols being released (i.e. captures a fraction *a* of the droplets) and the susceptibles wear a face mask with efficiency *b* (prevents a fraction *b* of the droplets from being inhaled) then Equation (2) becomes

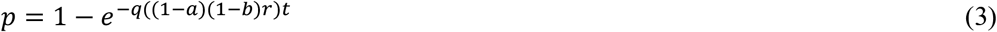

Furthermore, there may be periods where all the passengers within a cabin section remove their masks at the same time, for example during a meal service. Assuming a temporary removal of masks for a time duration, the infection probability becomes

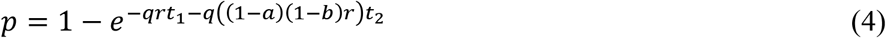

where *t*_1_ is the duration where none of the passengers wear masks and *t*_2_ is the duration where all passengers wear masks.

The model parameters and associated values used in the analysis are presented in Table 1. Three quanta generation rates are considered, 100 quanta/h, 20 quanta/h and 5.0 quanta/h, representing nominal Severe, Medium and Mild scenarios, respectively. These values are representative of the wide range of quanta generation rates suggested or derived from data for various COVID-19 transmission events reported in the literature [13, 18-20], (see Supplementary Table S1). These values are also consistent with quanta rate distributions (for breathing, speaking^*^, and speaking loudly^*^ index cases (^*^adjusted for resting conditions, see Supplementary Table S2) proposed by Buonanno et al. [18]. The mild, medium and severe cases lies between the 90^th^ to 95^th^ percentile of their breathing, speaking and speaking loudly distributions respectively. Further evidence justifying the use of the quanta release rate used in the severe case is derived from a known inflight COVID-19 asymptotic transmission event [4]. Using Equation (1) with the known infection probability derived for this inflight COVID-19 transmission event and the measured average aerosol reduction rate of 99.99% [14, 15], the quanta generation rate is back-calculated to be at least 102 quanta/h (see Supplementary Table S3). It is also noted that Marcus et al. [13] used 100 quanta/h as the high emission rate for aircraft COVID-19 infection risk analysis.

**Table 1.**
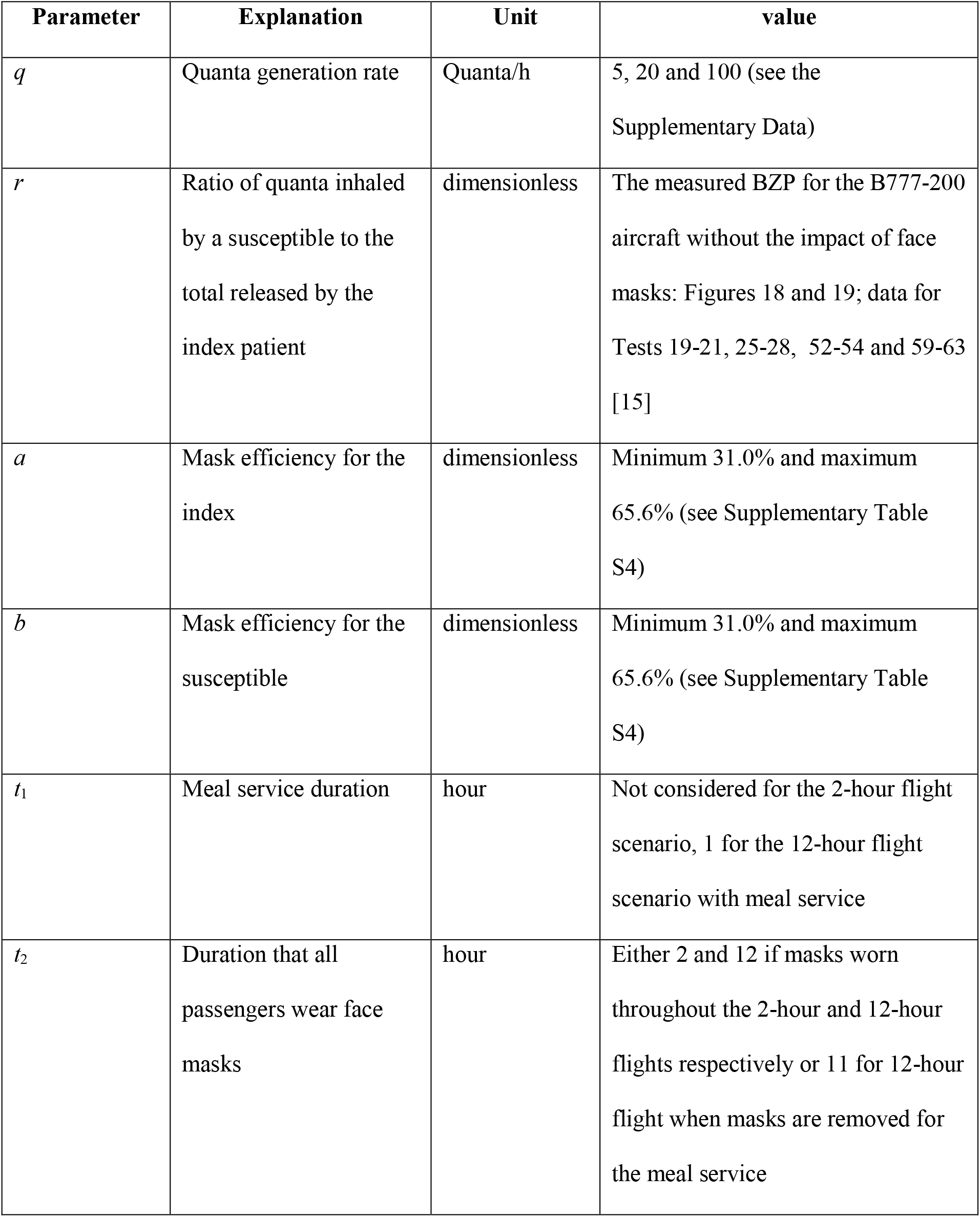
Model parameter and values.

| Parameter | Explanation | Unit | value |
| --- | --- | --- | --- |
| $q$ | Quanta generation rate | Quanta/h | 5, 20 and 100 (see the Supplementary Data) |
| $r$ | Ratio of quanta inhaled by a susceptible to the total released by the index patient | dimensionless | The measured BZP for the B777-200 aircraft without the impact of face masks: Figures 18 and 19; data for Tests 19-21, 25-28, 52-54 and 59-63 [15] |
| $a$ | Mask efficiency for the index | dimensionless | Minimum 31.0% and maximum 65.6% (see Supplementary Table S4) |
| $b$ | Mask efficiency for the susceptible | dimensionless | Minimum 31.0% and maximum 65.6% (see Supplementary Table S4) |
| $t_1$ | Meal service duration | hour | Not considered for the 2-hour flight scenario, 1 for the 12-hour flight scenario with meal service |
| $t_2$ | Duration that all passengers wear face masks | hour | Either 2 and 12 if masks worn throughout the 2-hour and 12-hour flights respectively or 11 for 12-hour flight when masks are removed for the meal service |

It is noted that the experimental BZP data (parameter *r* in Equation (3)) presented in [14, 15] should be scaled by a factor of 2.14 as the experiments used a sampling rate of 3.5 l/minute but the average respiratory rate for a resting human is 7.5 l/minutes. Two mask efficiencies of 31.0% and 65.6% are considered, derived from inflight aerosol experiment [14, 15] within the B777-200 cabin (see Supplementary Table S4). The infection probabilities derived in this analysis were determined for short- and long-haul flight-times of 2- and 12-hour respectively.

## Results

For a 2-hour flight, the MID-AFT cabin has a calculated maximum infection probability that varies from 4.5% (Mild Scenario) to 60.2% (Severe Scenario). The average infection probability varies from 0.1% to 2.5%. An average infection probability of 2.5% for the Severe Scenario implies that 1.2 of the 49 passengers seated in the monitored section of the cabin are likely to be infected. If the average infection probability applied to the entire cabin section, then 1.9 of the 75 passengers are likely to be infected. In contrast, the section with the smallest maximum and average probabilities of infection (FWD section) has a calculated maximum infection probability of 10.9% and an average infection probability of 0.9% (both for the Severe Scenario). With an average infection probability of 0.9%, if the average infection probability applied to the entire cabin section, then 0.4 of the 50 passengers and crew seated in the FWD section are likely to be infected.

Infection probability increases with exposure time however, the relative increase of infection probability within low risk locations is larger than those in high risk locations (see Table 2).

**Table 2.**
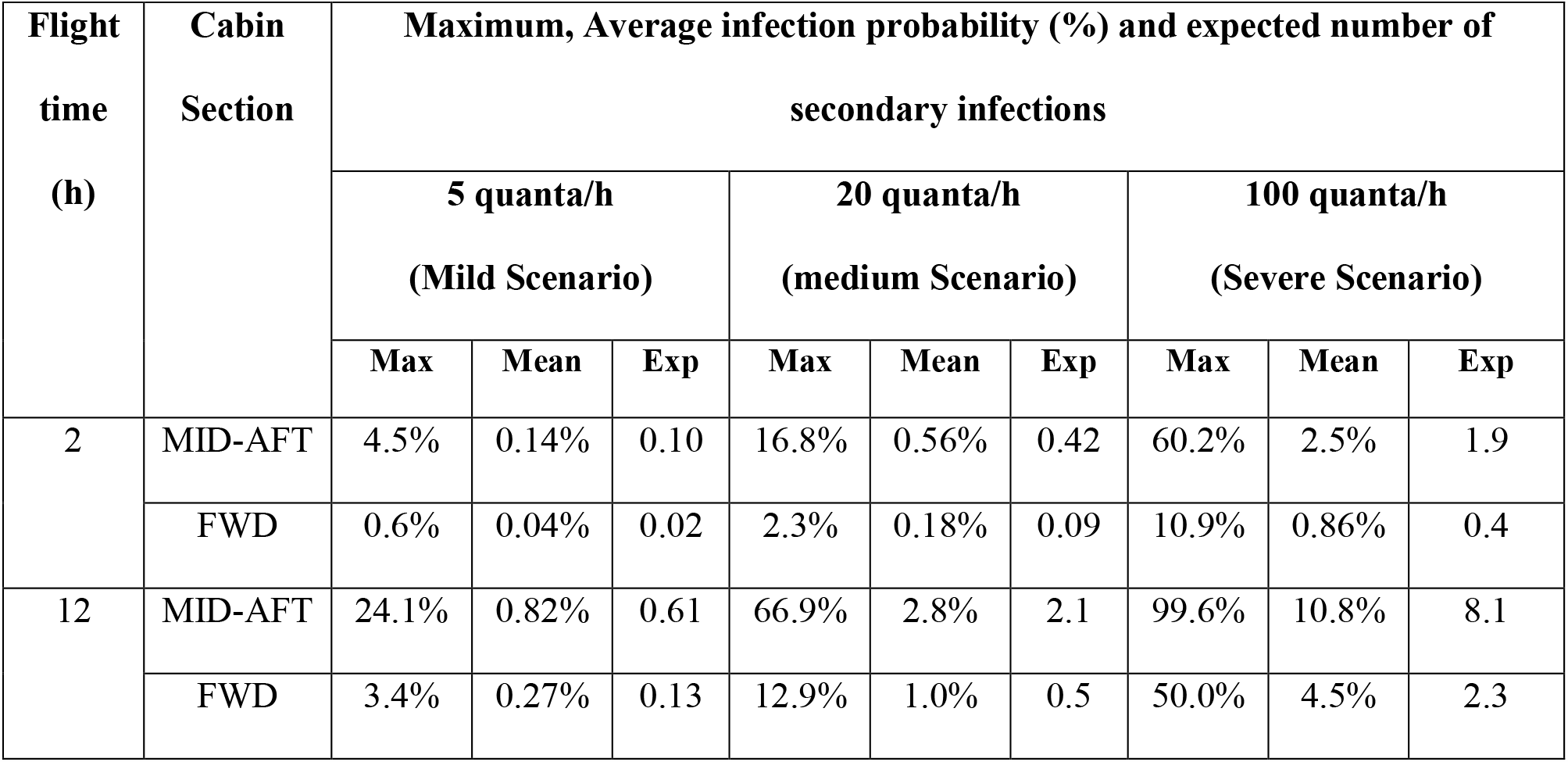
The estimated inflight infection probability and expected number of secondary infections without masks for 2-hour and 12-hour flights.

For a 12-hour flight, the estimated maximum infection probability (MID-AFT) varies from 24.1% (Mild Scenario) to 99.6% (Severe Scenario) while the average infection probability varies from 0.8% to 10.8%. With an average infection probability of 10.8% for the Severe Scenario, 5.3 of the 49 passengers seated in the monitored section of the cabin would be infected. If the average infection probability applied to the entire cabin section then 8.1 of the 75 passengers would be infected. In contrast, for the FWD section with an average infection probability of 4.5% (Severe Scenario), if the average infection probability applied to the entire cabin section, then 2.3 of the 50 passengers and crew would be infected.

The impact of wearing face coverings was next considered. As the MID-AFT section experiences the highest infection probability the mask analysis is focused on this section and is demonstrated for a 12-hour flight (see Table 3).

**Table 3.**
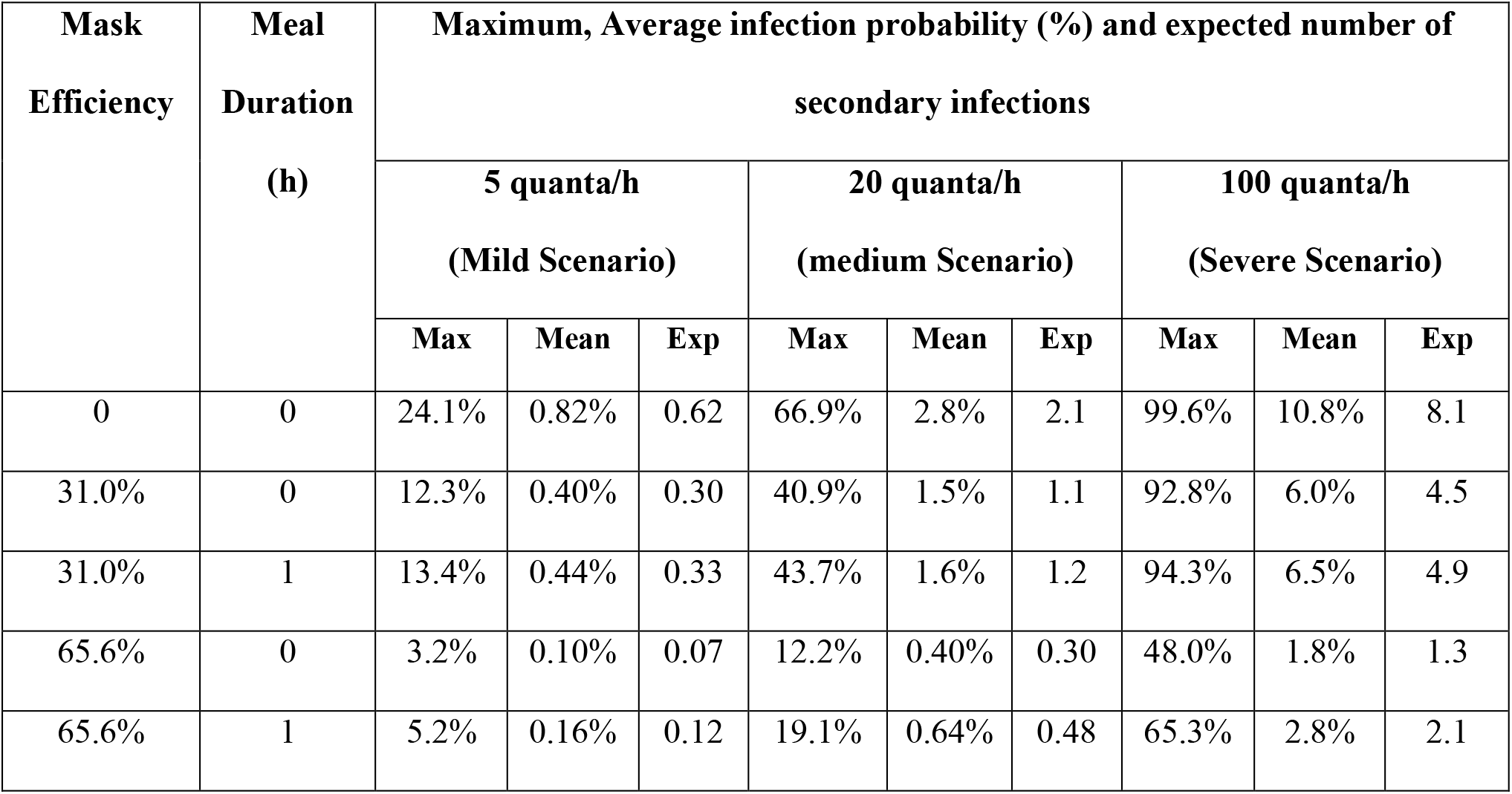
The estimated inflight infection probability and expected number of secondary infections for a 12-hour flight in the MID-AFT section

Assuming a mask effectiveness of 65.6%, the average infection probability is reduced from the range of 0.82% (Mild Scenario) to 10.8% (Severe Scenario) without masks to 0.10% to 1.8% with masks, an average reduction of 86% (see Table 3). In contrast, the maximum infection probability across the three scenarios is reduced by an average of 73%. With the less efficient masks, the average reduction in average infection probability is 47% while the average reduction in maximum infection probability is 32% (see Table 3). While even the low efficiency mask results in a substantial reduction in infection probability, there is still a high probability of infection with the high efficiency mask. With an average 1.8% probability of infection, 1.3 of the 75 passengers seated in the MID-AFT section are likely to be infected in the Severe Scenario even though they are wearing the high efficiency masks. It is also noted that the infection probability for a 2-hour flight without face masks (Table 2) are similar to those for a 12-hour flight with all passengers wearing highly effective face masks (Table 3).

On long duration flights, passengers are likely to remove their masks for some period of time, in particular during meal services. As meal services are usually undertaken at the same time within a given cabin section, most passengers will have their masks removed at the same time as others nearby, so increasing the probability of infection. For illustrative purposes we assume passengers spend a cumulative time of one hour for their meal with pre and post dinner drinks during their 12-hour flight.

When masks are worn but removed by all passengers for a 1-hour meal service, the average infection probability compared to the no mask scenario is reduced by an average of 77% and 43% across the three scenarios for the high and low efficiency masks respectively (see Table 3). The maximum infection probability across the three scenarios is reduced by an average of 61% and 28% for the high and low efficiency masks respectively. While this still represents a substantial improvement compared to the no mask situation, removing the high efficiency mask for a 1-hour period results in a 52%/59% increase in the maximum/average probability of infection compared to the situation where the mask is not removed. The increase in infection probability for low efficiency mask is significantly smaller at 6%/8%.

## Discussion

The key aspect of this work is the use of infection probability rather than time to infection to assess inflight transmission risk. Marcus et al. [13] estimated the time to infection of 7.5 hours for passengers without wearing masks and 34 hours with wearing face masks for the severe scenario with a quanta generation rate of 100 quanta/h. They stated that their model results were in general agreement with the modelling results from aircraft manufacturers [9-11] and with the empirical evidence from the US TRANSCOM study [14, 15] and so ‘*the estimated dose inhaled by an adjacent passenger over a few hours of exposure is likely to be less than the amount necessary to cause a secondary infection’*. With an identical quanta generation rate (100 quanta/h) and the maximum BZP data from the MID-AFT section [14, 15] we determine that the minimum time to infection (based on inhalation of 1 quanta) is 2.2 hours without masks, and using the low/high efficiency masks, the minimum time to infection becomes 4.6 hours/18.5 hours (see Figure 1). These times to infection are similar to those estimated in [13] and much less than those reported in [14].

**Figure 1:**
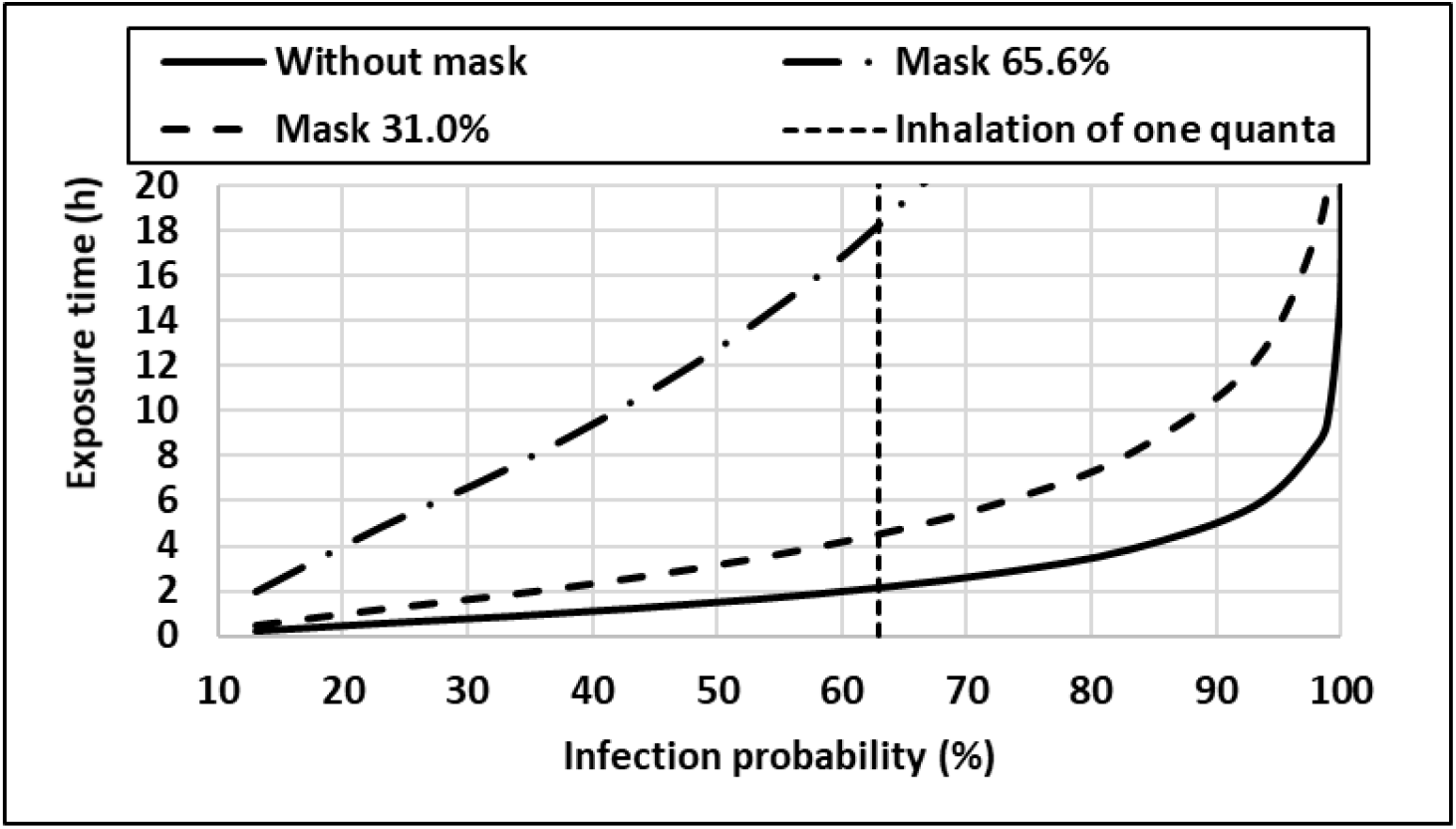
Exposure time and infection probability for the maximum BZP data in the MID-AFT section for the Severe Scenario

Nevertheless, we prefer to focus on the probabilities of infection within a given timeframe as some readers may misinterpret the “time to infection” to mean there is no possibility of infection if they are on-board for less than the stated timescales. If time to infection is used to define an end-point, it is therefore also necessary to specify how this end-point is defined, in particular the quanta dose required to cause infection or the corresponding infection probability associated with the end-point (see Figure 1). In our analysis of ‘time to infection’, we associate the inhalation of 1 quanta i.e. a 63% probability of being infected, with the end-point ‘being infected’. However, there is still a chance of being infected and hence a time for infection for shorter exposures. Furthermore, “time to infection” does not give an idea of the expected number of infections that could be caused by an index patient.

For the most extreme case considered, involving a 12-hour flight in which the passengers were not wearing masks, and for the Severe Scenario, the average probability of infection within the highest risk zone (MID-AFT) is 10.8%. With such a relatively low probability of infection it may be considered unlikely that an individual would be infected, unless they were in the seat of maximum exposure, in which case they would have a 99.6% probability of being infected. Furthermore, if all the passengers wore low efficiency masks for the entire 12-hour flight, this reduces the average probability of infection to 6.5% and this is further reduced to 2.8% if all the passengers are wearing high efficiency masks.

However, if these probabilities are applied to all the passengers (75) in the MID-AFT section, then the expected number of secondary infected cases is 8.1 with no masks and 1.3 when high efficiency masks are used. For the Medium and Mild scenarios the expected number of secondary infections is 2.1 and 0.6 respectively without masks and 0.3 and 0.1 respectively with high efficiency masks. This analysis suggests that multiple secondary infections can occur on-board aircraft, even though the passengers are protected with high ventilation rates and HEPA filters, although mask wearing helps to mitigate this risk. On average, the high/low efficiency masks reduce the average infection probability by 86%/47% and the maximum infection probability by 73%/32%.

Thus the wearing of masks by all passengers and crew should be considered an essential requirement for all flights and should be worn by all who can wear them at all times. However, even with the wearing of masks there is still a non-negligible probability of infection. Furthermore, removing masks, even for short periods on a 12-hour flight, such as for a 1-hour meal service, increases the average probability of infection by as much as 59%, compared to the situation where the mask is worn continuously.

In the Severe Scenario with high efficiency masks, removing the mask for 1-hour results in 2.1 of the 75 passengers seated in the MID-AFT section being infected, an increase of 0.8 passengers compared to the case where masks are worn throughout. If it is necessary to provide a meal service, airlines should keep the meal time to a minimum and consider alternating the meal times so that only half the passengers within a seat row are fed at any one time. In this way, the passengers either side of the passenger currently being fed are wearing masks, thus reducing the impact on the maximum infection probability associated with the passenger adjacent to the index passenger. Clearly, this will increase the time required for the meal service but will reduce the maximum infection probability.

There is also a clear class difference in the infection probabilities with the FWD (business class) section having much lower infection probabilities than the MID-AFT (economy class) section, which has the highest. The FWD average infection probability is about one third for the Mild and Medium scenarios and 42% for the severe scenario of the MID-AFT probabilities. These differences are probably due to a number of factors such as lower passenger numbers resulting in greater seating separations and the business class seat geometry possibly offering greater shielding. It is also not clear if the ventilation rate in the business class cabin section is greater than that in the economy class section.

The main limitation of this work is the assumed quanta generation rates because a reliable quanta generation rate distribution for COVID-19 is not known. However, the selected quanta generation rates used in the analysis are representative a wide range of quanta generation rates suggested from or derived from data for several COVID-19 transmission events reported in literature (see Supplementary Data). While the quanta generation categories appear representative, the proportion of passengers likely to be represented by each category is currently unknown. Furthermore, the emission rate of quanta from a given source is likely to vary during the flight as the index patient occasionally engages in speech rather than simply respiring and this is likely to be dependent on the nature of the passenger e.g. family groups, vacationers, business traveller. The model input data derived from the TRANSCOM experiments [14, 15] also has a number of limitations. Turbulent wakes produced by crew and passengers walking along the aisles were not considered. Furthermore, if the moving person is the index patient this is likely to spread infected aerosols over a larger portion of the cabin, possibly the entire aircraft if the infected person is a crew member. Finally, it is unlikely that both the index and susceptible passengers remain rigidly still within their seats during the entire duration of the flight, as assumed in the experiment.

## Data Availability

The data on which the analysis is based is available online in the third party pdf that can be obtained from the link provided.

https://www.usTRANSCOM.mil/cmd/docs/TRANSCOM%20Report%20Final.pdf

## Author’s contributions

ERG, ZW and AG conceived the idea, ZW and AG conducted the data analysis, ZW and ERG wrote the first draft and ZW, ERG, AG, JE, FJ worked collaboratively to edit and review the manuscript.

