## supplementary for "Inflight Transmission of COVID-19 Based on Aerosol Dispersion Data"

**Fire Safety Engineering Group**

**Faculty of Liberal Arts and Sciences**

**University of Greenwich**

**London UK**

### S1.0 Quanta generation rates for COVID-19

Within the analysis presented in the paper [S1], the infection probability for the COVID-19 disease is dependent on the quanta generation rate. Three infection quanta generation rates are considered in [S1] to represent a range of possible infection scenarios. These are intended to represent a nominal mild infection scenario in which the quanta generation rate is 5.0 quanta/h, a nominal medium infection scenario with quanta generation rate of 20 quanta/h and a nominal severe infection scenario with quanta generation rate of 100 quanta/h. In this section we provide evidence to support the selection of these quanta generation rates to represent these three broad categories of infection.

#### S1.1 COVID-19 quanta generation rates reported in the literature

The quanta generation rate is thought to be dependent on the degree of infection of the index patient and the nature of the activity they are involved in, e.g. breathing while resting, breathing while involved in light/heavy activity, vocalisation volume, etc. As a result, a wide variety of quanta generation rates are suggested or derived from data for various COVID-19 transmission events reported in the literature [S2-S8] varying from the order $10^0$ to $10^2$ (see Table S1).

The three values selected for analysis in [S1] are representative of the wide range of quanta generation rates suggested in Table S1.

Table S1. Quanta generation rates reported in the literature.

| **Case** | **Quanta generation rate (quanta/h)** | **Notes and references** |
|---|---|---|
| 1 | 0.225 | Estimated in [S5] for events involving health care workers |
| 2 | 0.37 | Oral breathing condition during resting [S2] |
| 3 | <1 | Resting [S3] |
| 4 | 2.5 | Oral breathing condition during heavy activity [S2] |
| 5 | 5.0 | Vocalization [S2] |
| 6 | 32.0 | Singing/speaking loudly [S2] |
| 7 | 14-48 | Derived from reproductive numbers for COVID-19 transmission events [S4] |
| 8 | >100 | Asymptomatic infectious subject performing vocalisation during light activities [S3] |
| 9 | 14, 48, 100 | These three values are suggested for COVID-19 transmission assessment in [S6] for assessing inflight events. They cover the range from lower to high generation rates. |
| 10 | 970±390 | Estimated in [S8] from a superspreading Choir event |

### S1.2 COVID-19 quanta generation rate distribution for people at rest

Buonanno et al. [S2] defined a number of quanta emission rate frequencies. This included breathing (resting), speaking (light activity), singing/speaking loudly (light activity). For inflight infection scenarios it is assumed that aircraft passengers will be seated for the vast majority of the flight and can therefore be considered to have a resting breathing rate. The speaking and speaking loudly emission rates were therefore adjusted for a resting breathing rate by multiplying by a scaling factor of 0.3551 (resting breathing rate (0.49 $m^3$/h) divided by the light activity breathing rate (1.38 $m^3$/h)). The adjusted rates are presented in Table S2.

Table S2. The percentile distribution of quanta emission rates (quanta/h) for breathing, speaking, and singing as defined by Buonanno et al. and adjustments for speaking distributions while at rest.

| Quanta emission rate type | $5^{th}$ | $25^{th}$ | $50^{th}$ | $75^{th}$ | $90^{th}$ | $95^{th}$ | $99^{th}$ |
|---|---|---|---|---|---|---|---|
| Breathing | 0.024 | 0.12 | 0.37 | 1.1 | 3.1 | 5.7 | 17 |
| Speak(original) | 0.32 | 1.6 | 5.0 | 15 | 42 | 76 | 240 |
| Speaking* | 0.11 | 0.57 | 1.8 | 5.3 | 14 | 26 | 85 |
| Singing/speaking loudly (original) | 2.1l | 10 | 32 | 98 | 270 | 490 | 1500 |
| Speaking* (loud) | 0.75 | 3.6 | 11.4 | 34.8 | 95.9 | 174 | 532 |

*the original values were based on light activity, these have been adjusted for a resting breathing rate in [S1].

The mild quanta release rate (5 quanta/h) used in [S1] lies between the $90^{th}$ and $95^{th}$ percentile of the breathing quanta emission distribution. Similarly, the medium quanta release rate (20 quanta/h) lies between the $90^{th}$ and $95^{th}$ percentile of the adjusted speaking distribution and the severe quanta release rate (100 quanta/h) lies between the $90^{th}$ and $95^{th}$ percentile of the adjusted speaking loudly distribution. This suggests that the quanta generation rates used to represent the three scenarios for analysis in [S1] are representative of the upper end of reported values and so are considered conservative.

**S1.3 COVID-19 quanta generation rates for inflight super spreading events**

Research into documented outbreaks using extreme value theory suggests that, for SARS-CoV-2, the distribution of secondary cases is consistent with being fat-tailed, implying that large super spreading events although rare, make a significant contribution in the overall transmission of the disease [S12]. Two inflight COVID-19 super spreading events have been reported in the literature [S7, S9]. In one asymptomatic transmission event [S7], seven passengers who arrived in New Zealand on 29 September 2020 on flight EK448 from United Arab Emirates were confirmed as COVID-19 cases. These passengers originated from five different countries. Among this group, five including the two index patients, reported negative test results in their country of origin at most 72 hours prior to boarding the flight. An academic research study, involving disease progression, travel dynamics and genomic analysis, suggests that at least four in-flight transmission cases of SARS-CoV-2 are likely to have occurred caused by the two index patients travelling together [S7]. There were eight other passengers seated with these confirmed cases in five seat rows during the 18-hour flight. The two index patients reported taking off their masks when they slept and when they were seated in the cabin. The seat rows covering the locations of the two index patients and the secondary infections in the transmission event on flight EK448 [S7] are similar to that for the index and susceptibles in Test 19-21 in [S10, S11] (see Figure S1).

In order to estimate the quanta generation rate for this asymptomatic inflight transmission event, it is assumed that the air ventilation state in this event on flight EK448 (B777-300ER) is similar to that in B777-200 and B767-300 cabins in the aerosol experiments [S10, S11]. Using this assumption, the ratio of the number of inhaled quanta to the total released quanta is on average 0.01% according the to the reported average aerosol reduction of 99.99% in the experiments. Using the parameter values for this event presented in Table S3, the quanta generation rate for this event is back-calculated using Equation (2) from [S1], expressed as

$$q = -\frac{ln(1-p)}{Irt} \qquad \text{(S1)}$$

Using this approach suggests that the estimated quanta generation rate for this event is at least 102 quanta/h for each of the two index patients, resulting in at least four inflight transmission cases in this investigated event (likely case 1 in Table S3). If there was only one index patient or more inflight transmission cases (likely cases 2-5 in Table S3), the quanta generation rate would increase up to 311 quanta/h per index patient (likely case 5 in Table S3).

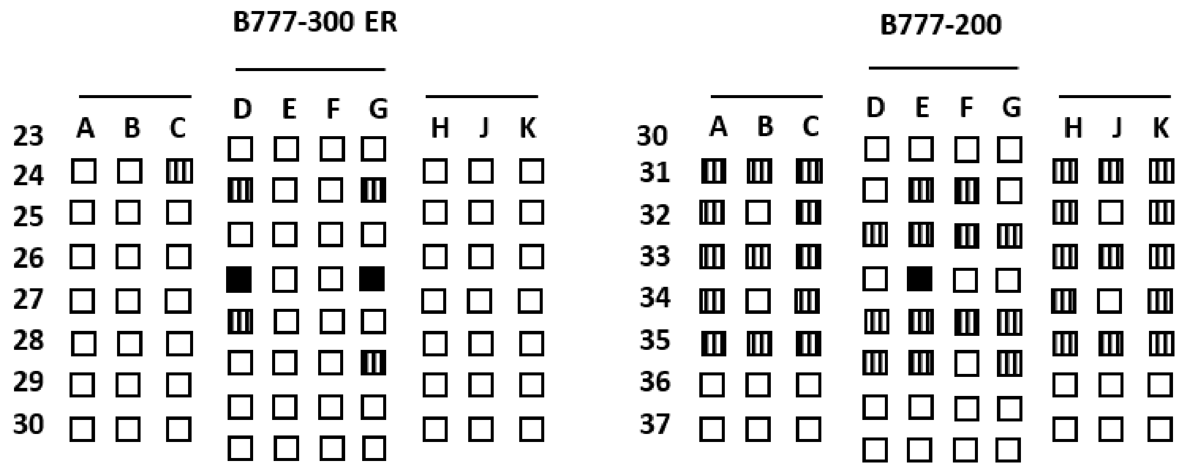


Figure S1. Locations of the index patients (solid fill) and the infections (lines) in flight EK448 transmission event [S7] (left) and the locations of aerosol source (solid fill) and sampling locations (lines) in the inflight experiments [S10, S11].

Table S3. Parameter values for flight EK448 asymptomatic transmission event and the estimated quanta generation rates with various combinations of indexes and infections.

| Likely case | Number of Index patients, *I* | Number of infections | Number of susceptibles | Infection probability, *p*, (%) | Flight time (h) | Inhaled quanta ratio (%) | Quanta generation rate per index patient (quanta/h) |
|---|---|---|---|---|---|---|---|
| 1 | 2 | 4 | 13 | 30.8 | 18 | 0.01 | 102 |
| 2 | 2 | 5 | 13 | 38.5 | 18 | 0.01 | 135 |
| 3 | 1 | 4 | 14 | 28.6 | 18 | 0.01 | 187 |
| 4 | 1 | 5 | 14 | 35.7 | 18 | 0.01 | 245 |
| 5 | 1 | 6 | 14 | 42.9 | 18 | 0.01 | 311 |

Using the same approach, we estimate that the quanta generation rate for the inflight transmission event in [S9] is 255 quanta/h.

This suggests that the quanta generation rate used to represent the Severe Scenario for analysis in [S1] is not an extreme value but representative of reported inflight super spreader events.

**S2.0 Mask efficiency derived from the Transcom aerosol dispersion experiments**

Within the analysis presented in the paper [S1], infection probability is determined for scenarios involving the use of masks. To simplify the analysis a mask filtration efficiency is used to represent the filtration of aerosols for both the source and susceptibles. Two mask efficiencies are used in [S1], 31% and 65.6%, intended to represent low and high mask efficiencies and are derived from the B777-200 aerosol experiments in [S10, S11].

In the aerosol dispersion experiments [S10, S11], several experiments were conducted involving the use of standard surgical masks where the mask was applied to the face of the mannequin representing the source (index patient). The modelled susceptibles did not wear face masks and so the effect of masks worn by index and susceptible was not directly measured in the experiments.

However, the mask efficiency associated with the index patient wearing the mask can be approximated from the measured breathing zone penetration data. This approach estimates the mask efficiency achieved in the experiments and so is specific to the conditions of the experiment including the adopted procedure used to represent the expired droplets, such as the exit flow rate of the particle chamber, whether the source used a continuous or discontinuous droplet release, how well the mask fitted the mannequin, etc.

The mask efficiency for the source is calculated by taking the difference in the breathing zone penetration data between the BNM (breathing source has no mask) and BM (breathing source has a mask) experiments and dividing the difference by the breathing zone penetration in the BNM experiment. These values are presented in Table S4 for each experiment pair (BNM and BM).

Table S4. Face mask efficiencies derived from B777-200 aerosol experiments in [S10, S11].

| **Experiment** | | **Breathing zone penetration BNM** | **Breathing zone penetration BM** | **Mask efficiency** |
|---|---|---|---|---|
| **Terminal** | **MID-AFT** | 0.082% | 0.050% | 39.0% |
| | **FWD-MID** | 0.012% | 0.008% | 33.3% |
| **Inflight** | **AFT** | 0.072% | 0.042% | 41.7% |
| | **MID-AFT** | 0.215% | 0.074% | 65.6% |
| | **FWD-MID** | 0.029% | 0.020% | 31.0% |
| | **FWD** | 0.027% | 0.013% | 51.9% |

As can be seen, the mask efficiency range for the surgical face masks used in these experiments varies from 31.0% to 65.6%. These filtration efficiencies appear to be within the range for surgical face masks reported in the literature, which are reported to be between 35% and 75% [S13], depending on the quality of the fit, with the highest efficiencies only for very good fit or when using sealed edge testing.

The calculated mask efficiency is for the index patient (source) however, within [S1] the same mask efficiency is used for both the index and susceptible. Generally, it is thought that the mask efficiency for the susceptible, when wearing the same mask as the index, is less than that for the index. If this is correct, the analysis in [S1] possibly over estimates the effectiveness of wearing masks.

**References**

[S1] Wang Z, Galea ER, Grandison A, Ewer Jand Jia F, Inflight Transmission Analysis of SARS-CoV-2 Based on Aerosol Dispersion Data, Submitted to Journal of Travel Medicine, December 2020.

[S2] Buonanno G, Morawska L, Stabile L, Quantitative assessment of the risk of airborne transmission of SARS-CoV-2 infection: Prospective and retrospective applications, *Environment International*, 2020, 145: https://doi.org/10.1016/j.envint.2020.106112

[S3] Buonanno G, Stabile L, Morawska L, Estimation of airborne viral emission: Quanta emission rate of SARS-CoV-2 for infection risk assessment, *Environment International*, 2020: 141.

[S4] Dai H, Zhao B, Association of the infection probability of COVID-19 with ventilation rates in confined spaces, *Build. Simulation*, 2020. https://doi.org/10.1007/s12273-020-0703-5

[S5] Hota B, Stein B, Lin M. et al. Estimate of airborne transmission of SARS-CoV-2 using real time tracking of health care workers. 2020. *medRxiv*: 2020.2007.2015.20154567

[S6] Marcus LJ, Assessment of Risks of SARS-CoV-2 Transmission During Air Travel and Non-Pharmaceutical Interventions to Reduce Risk, Phase One Report: Gate-to-Gate Travel Onboard Aircraft, Faculty and Scientists at the Harvard T.H. Chan School of Public Health. https://cdn1.sph.harvard.edu/wp-content/uploads/sites/2443/2020/10/HSPH-APHI-Phase-One-Report.pdf

[S7] Swadi T, Geoghegan JL, Devine T et al. A case study of extended in flight transmission of SARS CoV 2 en route to Aotearoa New Zealand, https://cdn.aerotelegraph.com/production/uploads/2020/11/A-case-study-of-extended-in-flight-transmission-of-SARS-CoV-2-en-route-to-Aotearoa-New-Zealand.pdf

[S8] Miller SL, Nazaroff WW, Jimenez JL et al. Transmission of SARS-CoV-2 by inhalation of respiratory, *Indoor Air*. 2020; DOI: 10.1111/ina.12751

[S9] Khanh NC, Thai PQ, Quach HL et al. Transmission of SARS-CoV 2 during long-haul flight, *Emerging Infectious Diseases*, 2020, 26: 2617-2624.

[S10] Silcott D, Kinahan S, Santarpia J et al, *TRANSCOM/AMC Commercial Aircraft Cabin Aerosol Dispersion Tests*, Submitted to: United States Transportation Command (USTRANSCOM) & Air Mobility Command (AMC), 2020. 15 Oct 2020.

[S11] Silcott D, Kinahan S, Santarpia J et al, *TRANSCOM/AMC Commercial Aircraft Cabin Aerosol Dispersion Tests*, Submitted to: United States Transportation Command (USTRANSCOM) & Air Mobility Command (AMC), 2020. 17 Nov 2020. https://www.usTRANSCOM.mil/cmd/docs/TRANSCOM%20Report%20Final.pdf

[S12] Wonga F, Collins JJ, Evidence that coronavirus superspreading is fat-tailed, *PNAS*, 2020, Vol. 117.

[S13] Rossettie S, Perry C, Pourghaed M, Zumwalt M, Effectiveness of manufactured surgical masks, respirators, and home-made masks in prevention of respiratory infection due to airborne microorganisms. https://pulmonarychronicles.com/index.php/pulmonarychronicles/article/view/675/1523
